# Evaluating a Locally Deployed 20-Billion Parameter Large Language Model for Automated Abstract Screening in Systematic Reviews

**DOI:** 10.64898/2026.03.04.26347506

**Authors:** Paulo Henrique Moreira Melo, Elena Guadagno, Dan Poenaru

**Affiliations:** Faculty of Medicine and Health Sciences, McGill University, Montreal, Quebec, Canada; Harvey E. Beardmore Division of Pediatric Surgery, The Montreal Children’s Hospital, McGill University Health Centre, Montreal, Quebec, Canada

**Keywords:** systematic review, large language model, abstract screening, artificial intelligence, natural language processing, evidence synthesis, local deployment

## Abstract

**Background:** Systematic reviews (SRs) are essential for evidence-based medicine but require extensive time and resources for abstract screening. Large language models (LLMs) offer potential for automating this process, yet concerns about data privacy, intellectual property protection, and reproducibility limit the use of cloud-based solutions in research settings.

**Objective:** To evaluate the performance of a locally deployed 20-billion parameter LLM for automated abstract screening in systematic reviews using a sensitivity-enhanced prompting strategy, with blind expert adjudication of all discordant human-AI cases.

**Methods:** We deployed GPT-OSS:20B locally using Ollama and evaluated its performance across three systematic reviews: AI applications in pediatric surgical pathology (n=3,350), LLM applications in electronic health records (n=4,326), and parental stress/caregiver burden in surgically treated children (n=8,970). A sensitivity-enhanced prompting strategy instructing the model to include abstracts when uncertain was employed. All discordant cases underwent blind expert adjudication.

**Results:** Across 16,646 abstracts, the LLM demonstrated variable sensitivity after expert adjudication: 100% in SR1, 95.7% in SR2, and 85.7% in SR3. Expert adjudication identified 11 human screening errors across all reviews that the LLM had correctly classified. The LLM completed screening 4.7 times faster than human reviewers.

**Conclusions:** A locally deployed LLM with sensitivity-enhanced prompting shows promising performance for systematic review abstract screening, particularly for technology-focused topics. Performance variability across domains suggests that screening accuracy depends partly on the objectivity of inclusion criteria. We recommend deploying LLMs as second screeners alongside human reviewers until performance is more fully validated across diverse domains.

## Introduction

Systematic reviews (SRs) are the cornerstone of evidence-based medicine. The Cochrane methodology requires at least two independent reviewers to screen all abstracts against predefined criteria, with a third expert adjudicating discordant cases [1,2]. For SRs with complex search strategies often identifying 5,000-10,000 abstracts, this process is profoundly resource-intensive, with abstract screening alone often requiring weeks of dedicated effort by multiple reviewers. The exponential growth of global research outputs has made it increasingly difficult for traditional literature review methods to keep pace, often rendering reviews outdated by the time of publication [3].
Large language models (LLMs) have emerged as promising tools for automating various aspects of systematic review workflows, including abstract screening [4-6,19]. Recent studies demonstrate that LLMs can achieve high sensitivity in screening tasks. Sanghera et al. evaluated multiple LLMs across 23 Cochrane reviews and found that LLM ensembles exhibited superior sensitivity compared to human reviewers (LLM max = 1.000 vs human max = 0.775), with workload reductions ranging from 37% to 99% [7,13]. Delgado-Chaves et al. demonstrated that LLM performance depends heavily on the interplay between inclusion/exclusion criteria and the model [8]. Guo et al. compared multiple LLM tools and found that ChatGPT v4.0 demonstrated accuracy consistently reaching or exceeding 90% [9,14,16].

Prompt engineering has proven critical for optimizing LLM screening performance. Gargari et al. demonstrated that including an “inclusivity sentence” instructing the model to include studies when uncertain elevated sensitivity from 62% to 95% [10,15,17]. This approach aligns with systematic review methodology: when an abstract is excluded, it is permanently removed from consideration; a false negative (FN) represents an irreversible error. In contrast, false positives (FP) simply advance to full-text screening where they can be identified and excluded.

Despite promising results, concerns about data privacy, reproducibility, and cost limit the adoption of cloud-based APIs in research settings [8,11,20]. Local deployment of open-source models addresses these barriers by ensuring data never leaves the institutional environment and maintaining complete version control [12]. The present study evaluates a locally deployed 20-billion parameter LLM for abstract screening across three systematic reviews using explicit sensitivity-enhanced prompting (“When in doubt, INCLUDE”). We validate performance through blind expert adjudication of all discordant cases, enabling quantification of errors in both human and AI screening.

## Methods

### Study Design and Data Sources

This methodological study evaluated the performance of a locally deployed LLM for systematic review abstract screening across three independent systematic reviews. Three systematic reviews were included: SR1 examined AI applications in pediatric surgical pathology (3,350 abstracts); SR2 examined LLM applications in electronic health records (4,326 abstracts); SR3 examined parental stress and caregiver burden in surgically treated children (8,970 abstracts). A total of 16,646 abstracts were screened.

### Technical Configuration and Prompting Strategy

We deployed GPT-OSS:20B, a 20-billion parameter open-source language model, locally using the Ollama framework [12]. This configuration ensures complete data privacy and guarantees version control and reproducibility. The input for each screening decision consisted of the article title, abstract, and the systematic review’s inclusion and exclusion criteria. The model output was structured as a JSON object containing a binary decision (include/exclude) and reasoning.

We employed a sensitivity-enhanced prompting strategy that instructed the model to include abstracts when uncertain. The rationale is that excluding a relevant study (FN) represents a more serious error than including an irrelevant study (FP), since excluded abstracts are never reviewed while included abstracts undergo subsequent human evaluation [7,10]. The prompt explicitly stated: “When in doubt, INCLUDE the abstract for further review.”

### Expert Adjudication and Performance Metrics

All discordant cases (where LLM and human reviewer disagreed) underwent blind expert adjudication by a senior researcher with extensive systematic review experience. The expert was blinded to which decision was from the LLM or human. Performance was assessed using sensitivity, specificity, counts of FN and FP, and human errors (cases where expert sided with LLM against original human decision).

## Results

Table 1 presents the initial screening results before expert adjudication. Prior to expert review, the LLM had 20 apparent FN and 559 FP across all reviews.

**Table 1.** Initial LLM screening performance (before expert adjudication)

| Systematic Review | Abstracts | Human Included* | AI Included | FN | FP |
| --- | --- | --- | --- | --- | --- |
| SR1: AI in Pediatric Surgery | 3,350 | 31 | 102 | 1 | 72 |
| SR2: LLMs in EHR | 4,326 | 49 | 534 | 5 | 486 |
| SR3: Parental Stress/Caregiver Burden | 8,970 | 79 | 66 | 14 | 1 |
| **Total** | **16,646** | **159** | **702** | **20** | **559** |

\*Original human reviewer decisions, before expert adjudication. FN = False Negatives; FP = False Positives.

Expert adjudication revised these findings (Table 2). In SR1, the one apparent FN was adjudicated as a correct LLM exclusion; 3 of 72 FPs were relevant studies that humans had incorrectly excluded (human errors: 3). In SR2, 3 of 5 FNs were adjudicated as correct LLM exclusions, leaving 2 true FN; 1 of 486 FPs was confirmed as a correct LLM inclusion (human errors: 4). In SR3, 3 of 14 FNs were adjudicated as correct LLM exclusions, leaving 11 true FN; the 1 FP was confirmed as a correct LLM inclusion (human errors: four).

**Table 2.** Final LLM screening performance after expert adjudication.

| Systematic Review | True Rel. | Sensitivity | Specificity | FN | FP | Human Err |
| --- | --- | --- | --- | --- | --- | --- |
| SR1: AI in Pediatric Surgery | 33 | 100% | 97.9% | 0 | 69 | 3 |
| SR2: LLMs in EHR | 47 | 95.7% | 88.7% | 2 | 485 | 4 |
| SR3: Parental Stress/Caregiver | 77 | 85.7% | 99.99% | 11 | 0 | 4 |
| **Overall** | **157** | **91.7%** | **96.7%** | **13** | **554** | **11** |

True Rel. = True Relevant per expert adjudication. Human Err = cases where expert sided with LLM against original human decision.

The LLM completed screening of the 3,350 abstracts in SR1 in 5.58 hours, compared to 26 hours required by human reviewers, representing a 4.7-fold improvement in efficiency. On average, the LLM processed approximately six seconds per abstract.

## Discussion

This study demonstrates that a locally deployed LLM with sensitivity-enhanced prompting can achieve high but variable performance for systematic review abstract screening. The model achieved excellent sensitivity for technology-focused reviews (100% SR1, 95.7% SR2) but lower sensitivity for psychosocial outcomes (85.7% SR3). This variability likely reflects differences in the objectivity of inclusion criteria: SR1/SR2 involved concrete, well-defined technology criteria, whereas SR3’s focus on parental stress and caregiver burden left more room for interpretation. This is consistent with Delgado-Chaves et al., who found that clearer criteria lead to better LLM performance [8,14]. Systematic reviews in social sciences may be more challenging due to subjective criteria.

A key finding was bidirectional misclassification by humans and the LLM. The LLM caught 11 relevant studies that humans had wrongly excluded, while humans caught 13 that the LLM missed. This suggests the two approaches complement each other; combining them likely works better than relying on either alone [13,16]. Expert adjudication proved essential here; without it, we would have treated human decisions as the gold standard and drawn wrong conclusions about how well the LLM actually performed.

Based on these findings, we recommend using LLMs as second screeners alongside human reviewers until their sensitivity is better validated across different topics. In practice, a human screens all abstracts, the LLM screens independently, and disagreements go to expert adjudication. This keeps the rigor of dual screening while cutting down on personnel [18,19]. Local deployment also addresses key adoption barriers like data privacy and reproducibility.

This study has limitations worth noting. Performance varied considerably by topic; the LLM excelled with technology-focused reviews but struggled more with psychosocial outcomes, so domain-specific validation is needed before deployment in new areas. The model gives binary yes/no outputs without confidence scores, which means it is not possible to easily flag borderline cases that might benefit from closer human attention. We only tested abstract screening, not full-text review, so it was not possible to determine how the LLM would perform at that stage. Additionally, all three reviews came from our research team; external validation across different institutions and reviewer styles would strengthen these findings.Finally, the 13 missed studies across SR2 and SR3 are a real limitation. These represent potentially important evidence that would have been lost without human involvement, reinforcing why human oversight remains necessary.

## Conclusion

Locally deployed LLMs with sensitivity-enhanced prompting show promising screening performance, achieving excellent results for technology-focused topics with more variable performance for psychosocial outcomes. Both LLMs and humans make errors, often on different abstracts, supporting human-AI collaboration rather than full automation. We recommend deploying LLMs as second screeners with expert adjudication until performance is validated across domains. Future work should develop confidence-calibrated outputs, validate across additional domains, and establish standardized benchmarks for LLM screening evaluation [19,21].

## Conflicts of Interest

The authors have no conflicts of interest to disclose.

## Data Availability

The datasets used in this study are available from the corresponding author upon reasonable request.

## Code Availability

The code for the LLM screening pipeline is publicly available at https://github.com/PauloHenriqueMelo/Systematic_Review_Screening_LLM

## Notes

### Competing Interest Statement

The authors have declared no competing interest.

### Funding Statement

This study did not receive any funding

